# Initial Experience in Predicting the Risk of Hospitalization of 496 Outpatients with COVID-19 Using a Telemedicine Risk Assessment Tool

**DOI:** 10.1101/2020.07.21.20159384

**Authors:** James B. O’Keefe, Elizabeth J. Tong, Thomas H. Taylor, Ghazala A. Datoo O’Keefe, David C. Tong

## Abstract

**Title:** Initial Experience in Validation of a Telemedicine Risk Assessment Tool for Hospitalization of Patients with COVID-19

**Importance:** Risk assessment of patients with acute coronavirus disease 2019 (COVID-19) in a telemedicine context is not well described. In the setting of large numbers of patients, a risk assessment tool may guide resource allocation not only for patient care but also for maximum healthcare and public health benefit.

**Objective:** To determine whether a risk prediction tool developed and implemented in March 2020 (“COVID-19 Telemedicine Risk Tier Assessment”) accurately predicts subsequent hospitalizations.

**Design:** Retrospective cohort study, enrollment from March 24 to May 26, 2020 with follow-up calls until hospitalization or clinical improvement (final call date range March 27 to June 19, 2020)

**Setting:** Single center “Virtual Outpatient Management Clinic” (VOMC) assessing and managing outpatients from a large quaternary medical system in Atlanta, Georgia

**Participants:** 496 patients with COVID-19 confirmed by nasopharyngeal sampling in isolation at home at the time of diagnosis. Exclusion criteria included: (1) hospitalization prior to VOMC enrollment, (2) immediate discharge from VOMC with no follow-up calls due to resolution.

**Exposure:** Acute COVID-19 illness

**Main Outcome and Measures:** Hospitalization was the outcome. Days to hospitalization was the metric. Survival analysis using Cox regression was used to determine factors associated with hospitalization.

**Results:** The risk-assessment rubric assigned 496 outpatients to tiers as follows: Tier 1, 237 (47.8%); Tier 2, 185 (37.3%); Tier 3, 74 (14.9%). Subsequent hospitalizations numbered 3 (1%), 15 (7%), and 17 (23%) and for Tiers 1-3, respectively. From a Cox regression model with age ≥ 60, gender, and self-reported obesity as covariates, the adjusted hazard ratios using Tier 1 as reference were: Tier 2 HR=3.74 (95% CI, 1.06-13.27; P=0.041); Tier 3 HR=10.87 (95% CI, 3.09-38.27; P<0.001). Tier was the strongest predictor of time to hospitalization.

**Conclusions and Relevance:** A telemedicine risk assessment tool prospectively applied to an outpatient population with COVID-19 identified both low-risk (Tier 1) and high-risk (Tier 3) patients with better performance than individual risk factors alone. This approach may be appropriate for optimum allocation of resources.

## Introduction

The severity of coronavirus disease 2019 (COVID-19) varies from asymptomatic to life-threatening.^1^,^2^ At the time of diagnosis, most patients have mild illness and do not require hospitalization.^3^ For these patients, the recommendation is to isolate at home and monitor symptoms under the care of a medical provider.^4^,^5^ Many U.S. medical centers have employed telemedicine and remote monitoring programs to provide this care.^6^-^8^ Monitoring programs require investment and staffing;^7^ it may be appropriate to focus these resources on those at highest risk of hospitalization for severe COVID-19. While it is recognized that certain groups (e.g. older adults, patients with diabetes)^9^-^12^ have higher rates of hospitalization, there are no validated risk assessment tools that stratify risk for outpatients undergoing home monitoring.^13^ The tools in existence often require in-person criteria (e.g. vital signs, labs, imaging) not available by telemedicine.^13^,^14^

In order to better target care for outpatients with COVID-19, we created a risk assessment tool based on published data available in March 2020 (eAppendix 1). This tool incorporates age, comorbidities, symptom severity and course, and the ability to isolate – criteria highlighted in initial Centers for Disease Control (CDC) guidance for home monitoring of patients with COVID-19.^4^ Based on these factors, patients are assigned to a “risk tier 1-3” (indicating low, moderate, or high risk for hospitalization). For patients seen during acute illness, low-risk patients (Tier 1) must meet all of the following criteria: age <60, no comorbidities known to increase risk of severe COVID-19, no lower respiratory tract symptoms (except mild cough), able to self-isolate. High-risk patients (Tier 3) meet any of the following criteria: age ≥70, younger age with specific high-risk comorbidity or multiple comorbidities, new and/or worsening lower respiratory symptoms, persistent systemic symptoms (e.g. fevers >6 days), or uncertain ability to self-isolate. Exceptions to the criteria included: (1) provider discretion to override the risk assessment tool; (2) patients who appeared to be improving after the second week of illness could be assigned to a low tier even if older age or comorbidities were present.

We prospectively applied this risk assessment tool during the telemedicine assessment of outpatients diagnosed with COVID-19 in a large quaternary academic health system in Atlanta, GA. Patients were followed with regular phone calls (frequency and duration varied by risk tier) until clinical improvement or hospitalization. The clinical care pathway for outpatients with COVID-19 in our clinic, the Virtual Outpatient Monitoring Clinic (VOMC), is outlined in eAppendix 2. In this retrospective study, we analyzed patient data gathered systematically at VOMC intake visits, including patient characteristics and assigned risk tier, and used an outcome of hospitalization related to COVID-19. We hypothesized that the multifactorial tool would predict hospitalization rates. Below, we outline our findings and our initial experience using this risk assessment tool.

## Methods

### Ethics

The study was approved by the Emory University Institutional Review Board and the requirement for consent was waived as the study posed no more than minimal risk.

### Study setting and population

The study is a retrospective cohort investigation of outpatients with confirmed COVID-19 at Emory Healthcare, the largest academic health system in Georgia (serving the greater Atlanta metropolitan area). During the study period, outpatient testing was performed by nasopharyngeal sampling for real-time reverse transcription–polymerase chain reaction (RT-PCR) detection of severe acute respiratory syndrome coronavirus 2 (SARS-CoV-2). Adult patients with positive RT-PCR results from the screening clinics or emergency departments were called by a result notification team to provide self-care advice and refer for enrollment in the VOMC.

The VOMC comprised an intake team of 14 physicians and 3 advanced practice providers (APPs) from two primary care clinics; and follow-up call teams included 19 redeployed registered nurses (RNs) and 20 APPs. All intake providers were trained in the use of the risk assessment tool in a one-hour webinar and conducted a median of 25 intake visits during the study period [range: 5-99].

Enrollment criteria for this study included: (1) completion of new patient VOMC visit during the period of March 24 to May 26, 2020, and (2) Documentation of positive RT-PCR for SARS-CoV-2. Exclusion criteria include: (1) hospitalization prior to VOMC enrollment, (2) immediate discharge from VOMC (no follow-up calls) due to no care needs.

### Outcome

Hospitalization was the primary study outcome, consistent with stated purpose of the risk assessment tool. Emergency department visits and observation admissions were not included as events. Hospitalization at four Emory Healthcare acute care hospitals was determined by Emory Clinical Data Warehouse (CDW) queries, last performed July 6, 2020. External hospitalizations were identified by chart review in (1) clinical notes, (2) administrative messages; and (3) hospitalization documentation in the Emory electronic health record per data sharing agreements with other health systems. Loss to follow-up was minimal because VOMC patients were followed for minimum pre-specified periods with regular telephone calls and notifications of external hospitalizations were documented in the medical record. To validate this approach to identification of hospitalizations, we were able to compare our chart reviews of hospitalizations to patient-reported hospitalizations in a separate long-term follow-up telephone survey of 158 patients.

### Covariates

Risk assessment data were obtained for all patients enrolling in the VOMC during a scheduled telemedicine visit utilizing synchronous two-way audio/video communication (with telephone call as backup option). VOMC intake providers completed a standard note template, including demographics, comorbidities (past medical history and specific conditions with elevated COVID-19 severity risk), symptom description (onset, severity, and course), social support and ability to isolate, and clinician-assigned risk tier using the risk assessment tool (eAppendix 1). These data were extracted from the clinical notes by CDW query. Missing data were included, when possible, by manual chart review by the authors (JO, GO) of provider documentation in the intake note. Actual body mass index (BMI) was obtained with a second CDW query. In practice, the patients could change tiers to receive more or less frequent contact based on change in illness severity, but we used data collected at the initial telemedicine visit for analysis.

### Statistical methods

Survival analysis was used to determine factors associated with hospitalization to evaluate the risk tier model. Initial unadjusted hazard ratios (HRs) were calculated using a Cox proportional hazards model. A multivariable model was then constructed using a Cox proportional hazards model. Despite good assurance that we had good follow up in terms of hospitalizations we were conservative in our analysis and right censored patients at the last VOMC nurse phone call rather than extend the time variable to the end of the study. Time-varying covariates were identified by individual evaluation of covariates looking at Kaplan Meier curves and testing for a statistically significant time variable interaction. Covariates with a p value <0.05 for the interaction term were considered time-varying. Both forward stepwise likelihood ratio and backward likelihood ratio stepwise selection was performed. The models developed by backward and forward selection were then manually checked by adding and removing individual variables and assessing model fit. Pairwise deletion was used during the exploratory phase for any variables with missing data. The final model did not have any missing data. Logistic regression was also performed with the same variables to shadow the Cox regression analysis. All analyses were conducted using SPSS statistical software version 26 (IBM Corp).

## Results

### Participant characteristics

We identified 549 patients completing a new VOMC visit from March 24, 2020 through May 26, 2020 and included 496 patients in the analysis after excluding 7 patients without a positive RT-PCR, 26 patients hospitalized for COVID-19 prior to VOMC visit, and 20 patients who met criteria for discharge at the initial visit and were not placed into an initial Tier. Table 1 describes our patient population by tiers. At the initial visit, 237 patients (47.7%) were placed in Tier 1 (low-risk), 185 (37%) in Tier 2 (intermediate-risk), and 74 (14.9%) in Tier 3 (high-risk). The majority (330 patients, 66.5%) are female, 252 (50.8%) are black, and 383 (77.2%) were under age 60 years. Race was unknown or other for 147 (29.6%) of our patient population. Only 174 patients (35%) reported no high-risk comorbidities, with hypertension (175 patients, 35.3%) and reported BMI >30 (147 patients, 29.6%) as the most frequent comorbidities. Ability to self-isolate was documented as adequate for 409 patients (82.5%), inadequate for 9 patients (1.8%), and the rest were missing data. Most patients (316, 63.7%) had mild symptoms or no symptoms at the time of the visit. The mean age was 47.6 years; the mean days from first symptom to visit was 9.3, the mean days from RT-PCR test to visit was 3.7 days; the mean duration of follow-up telephone calls was 13.1 days.

**Table 1:** Characteristics of Outpatients with COVID-19 by Assigned Risk Tier

|  |  | All Patients | Tier |  |  |
| --- | --- | --- | --- | --- | --- |
|  |  | n=496 | 1 (n=237) | 2 (n=185) | 3 (n=74) |
| Mean (95% CI) |  |  |  |  |  |
| Age |  | 47.6<br>(46.3-48.9) | 41.5<br>(39.8-43.2) | 52.5<br>(50.6-54.4) | 54.9<br>(51.4-58.4) |
| Time from first symptom to Visit (Days) |  | 9.3 (8.5-10.0) | 8.9 (8.2-9.7) | 10.0 (8.4-11.6) | 8.4 (6.9-9.8) |
| Time from COVID test to Visit (Days) |  | 3.7 (3.4-3.9) | 3.9 (3.4-4.4) | 3.5 (3.0-3.9) | 3.3 (2.7-4.0) |
| Follow up duration (Days) |  | 13.1 (12.2-13.9) | 9.5 (8.6-10.4) | 16.3 (14.8-17.7) | 16.7 (14.0-19.3) |
| No. (%) |  |  |  |  |  |
| Age Category | 18-29 | 78 | 65 (83.3%) | 10 (12.8%) | 3 (3.8%) |
|  | 30-39 | 84 | 49 (58.3%) | 26 (31.0%) | 9 (10.7%) |
|  | 40-49 | 106 | 50 (47.2%) | 39 (36.8%) | 17 (16.0%) |
|  | 50-59 | 115 | 48 (41.7%) | 50 (43.5%) | 17 (14.8%) |
|  | 60-69 | 84 | 21 (25.0%) | 45 (53.6%) | 18 (21.4%) |
|  | ≥ 70 | 29 | 4 (13.8%) | 15 (51.7%) | 10 (34.5%) |
| Race | Black | 252 | 109 (43.3%) | 102 (40.5%) | 41 (16.3%) |
|  | White | 97 | 47 (48.5%) | 36 (37.1%) | 14 (14.4%) |
|  | Other | 147 | 81 (55.1%) | 47 (32.0%) | 19 (12.9%) |
| Gender | Female | 330 | 156 (47.3%) | 125 (37.9%) | 49 (14.8%) |
|  | Male | 166 | 81 (48.8%) | 60 (36.1%) | 25 (15.1%) |
| Comorbidities | Age ≥ 60 | 113 | 25 (22.1%) | 60 (53.1%) | 28 (24.8%) |
|  | Asthma | 73 | 18 (24.7%) | 37 (50.7%) | 18 (24.7%) |
|  | Cancer or malignancy | 37 | 9 (24.3%) | 21 (56.8%) | 7 (18.9%) |
|  | COPD | 5 | 0 (0.0%) | 4 (80.0%) | 1 (20.0%) |
|  | CAD | 24 | 1 (4.2%) | 11 (45.8%) | 12 (50.0%) |
|  | Diabetes | 69 | 10 (14.5%) | 35 (50.7%) | 24 (34.8%) |
|  | Drug abuse/addiction | 4 | 0 (0.0%) | 4 (100.0%) | 0 (0.0%) |
|  | Heart failure | 10 | 2 (20.0%) | 4 (40.0%) | 4 (40.0%) |
|  | Hypertension | 175 | 42 (24.0%) | 91 (52.0%) | 42 (24.0%) |
|  | Immune suppression | 30 | 9 (30.0%) | 11 (36.7%) | 10 (33.3%) |
|  | Lung disease | 17 | 3 (17.6%) | 9 (52.9%) | 5 (29.4%) |
|  | Reported obesity<sup>a</sup> | 147 | 52 (35.4%) | 63 (42.9%) | 32 (21.8%) |
|  | Corrected obesity | 212 | 87 (41.0%) | 85 (40.1%) | 40 (18.9%) |
|  | Renal disease | 16 | 3 (18.8%) | 6 (37.5%) | 7 (43.8%) |
| Number of Diagnoses | Healthy (0) | 174 | 129 (74.1%) | 35 (20.1%) | 10 (5.7%) |
|  | 1 | 158 | 81 (51.3%) | 65 (41.1%) | 12 (7.6%) |
|  | 2 | 91 | 19 (20.9%) | 48 (52.7%) | 24 (26.4%) |
|  | ≥ 3 | 73 | 8 (11.0%) | 37 (50.7%) | 28 (38.4%) |
| Ability to self isolate safely | Adequate | 409 | 205 (50.1%) | 156 (38.1%) | 48 (11.7%) |
|  | Inadequate | 9 | 1 (11.1%) | 3 (33.3%) | 5 (55.6%) |
|  | unknown | 78 | 31 (39.7%) | 26 (33.3%) | 21 (26.9%) |
| Severity of symptoms | none-mild | 316 | 204 (64.6%) | 102 (32.3%) | 10 (3.2%) |
|  | moderate | 134 | 18 (13.4%) | 69 (51.5%) | 47 (35.1%) |
|  | severe | 9 | 0 (0.0%) | 0 (0.0%) | 9 (100.0%) |
|  | unknown | 37 | 15 (40.5%) | 14 (37.8%) | 8 (21.6%) |
| Symptoms course | improving | 264 | 156 (59.1%) | 90 (34.1%) | 18 (6.8%) |
|  | stable | 155 | 61 (39.4%) | 65 (41.9%) | 29 (18.7%) |
|  | worsening | 31 | 0 (0.0%) | 14 (45.2%) | 17 (54.8%) |
|  | unknown | 46 | 20 (43.5%) | 16 (34.8%) | 10 (21.7%) |
| Abbreviations: COPD, chronic obstructive pulmonary disease; CAD, coronary artery disease |  |  |  |  |  |
| <sup>a</sup>Obesity defined as body mass index (BMI) greater than 30, calculated as weight in kilograms divided by height in meters squared |  |  |  |  |  |

### Univariate analysis

Table 2 demonstrates unadjusted hazard ratios, 95% confidence interval, and p value for each factor comparing 35 VOMC patients requiring hospitalization and 461 patients who did not require hospitalization during the follow-up period. Statistically significant factors included risk tier, age, coronary artery disease, diabetes mellitus, heart failure, reported obesity (BMI >30), 2 comorbidities, 3 or more comorbidities, severe symptom rating, and worsening symptom course. Of the patients initially categorized as Tier 3, 17 of 74 (23%) were hospitalized in the course of their care, compared with 15 of 185 (7%) for Tier 2 and 3 of 237 (1.3%) of Tier 1. Among 35 hospitalized patients, the median days to admission from symptom onset was 8 in Tier 3, 11 in Tier 2, and 13 in Tier 1. If we combined Tiers 2 and 3, the Tier model has a sensitivity of 91% and specificity of 51%. Likewise, if we combine Tiers 1 and 2 the Tier model has a sensitivity of 49% and a specificity of 88%. Tier had the highest unadjusted hazard ratio of all factors with 5.29 for Tier 2 and 16.24 for Tier 3 in comparison to Tier 1.

**Table 2:** Characteristics of Hospitalized and Non-hospitalized Patients with Outpatients with COVID-19

|  |  | Non-Hospitalized<br>(n=461) | Hospitalized<br>(n=35) | Unadjusted Hazard ratio<br>(95% CI, p value) |
| --- | --- | --- | --- | --- |
| (Mean, 95% CI) |  |  |  |  |
| Age |  | 46.7 (45.4-48.1) | 59.1 (55.2-63.1) | p<0.001 |
| Time from first symptom to Visit (Days) |  | 9.4 (8.6-10.2) | 7.4 (5.2-9.6) | p = 0.162 |
| Time from positive COVID to Visit (Days) |  | 3.7 (3.4-4.0) | 2.7 (1.9-3.5) | p = 0.088 |
| Follow up duration (Days) |  | 13.4 (12.6-14.3) | 8.5 (5.0-12.1) | p = 0.003 |
| No. (%) |  |  |  |  |
| Age Category | 18-29 | 78 (100.0%) | 0 (0.0%) | 0 |
|  | 30-39 | 81 (96.4%) | 3 (3.6%) | reference |
|  | 40-49 | 103 (97.2%) | 3 (2.8%) | 0.71 (0.14-3.53, 0.677) |
|  | 50-59 | 105 (91.3%) | 10 (8.7%) | 2.16 (0.59-7.85, 0.244) |
|  | 60-69 | 68 (81.0%) | 16 (19.0%) | 4.89 (1.42-16.79, 0.012) |
|  | ≥ 70 | 26 (89.7%) | 3 (10.3%) | 2.32 (0.47-11.52, 0.305) |
| Race | Black | 234 (92.9%) | 18 (7.1%) | reference |
|  | White | 86 (88.7%) | 11 (11.3%) | 1.63 (0.77-3.45, 0.202) |
|  | Other | 141 (95.9%) | 6 (4.1%) | 0.63 (0.25-1.59, 0.326) |
| Gender | Female | 311 (94.2%) | 19 (5.8%) | reference |
|  | Male | 150 (90.4%) | 16 (9.6%) | 1.76 (0.91-3.43, 0.095) |
| Comorbidities | Age ≥ 60 | 94 (83.2%) | 19 (16.8%) | 3.77 (1.94-7.34, <0.001) |
|  | Asthma | 67 (91.8%) | 6 (8.2%) | 1.07 (0.44-2.59, 0.877) |
|  | Cancer or malignancy | 34 (91.9%) | 3 (8.1%) | 1.07 (0.33-3.50, 0.908) |
|  | COPD | 4 (80.0%) | 1 (20.0%) | 2.58 (0.35-18.84, 0.352) |
|  | CAD | 18 (75.0%) | 6 (25.0%) | 3.71 (1.54-8.96, 0.004) |
|  | Diabetes | 56 (81.2%) | 13 (18.8%) | 3.59 (1.81-7.12, <0.001) |
|  | Drug abuse/addiction | 3 (75.0%) | 1 (25.0%) | 4.29 (0.59-31.45, 0.152) |
|  | Heart failure | 6 (60.0%) | 4 (40.0%) | 5.84 (2.06-16.55, 0.001) |
|  | Hypertension | 157 (89.7%) | 18 (10.3%) | 1.75 (0.90-3.40, 0.099) |
|  | Immune suppression | 30 (100.0%) | 0 (0.0%) | 0.05 (0.00-16.22, 0.302) |
|  | Lung disease | 14 (82.4%) | 3 (17.6%) | 2.10 (0.64-6.88, 0.221) |
|  | Obesity Reported | 130 (88.4%) | 17 (11.6%) | 2.27 (1.17-4.41, 0.015) |
|  | Obesity Corrected | 212 (815.4%) | 186 (715.4%) | 3.83 (1.80-8.18, 0.001) |
|  | Renal disease | 13 (81.3%) | 3 (18.8%) | 2.35 (0.72-7.71, 0.158) |
| Number of Diagnoses | Healthy (0) | 170 (97.7%) | 4 (2.3%) | reference |
|  | 1 | 148 (93.7%) | 10 (6.3%) | 2.61 (0.82-8.34, 0.105) |
|  | 2 | 83 (91.2%) | 8 (8.8%) | 3.43 (1.03-11.40, 0.044) |
|  | ≥ 3 | 60 (82.2%) | 13 (17.8%) | 6.77 (2.20-20.83, 0.001) |
| Ability to self isolate safely | Adequate | 383 (93.6%) | 26 (6.4%) | 0.26 (0.06-1.11, 0.069) |
|  | Inadequate | 7 (77.8%) | 2 (22.2%) | reference |
|  | unknown | 71 (91.0%) | 7 (9.0%) | unknown |
| Severity of symptoms | none-mild | 301 (95.3%) | 15 (4.7%) | reference |
|  | moderate | 121 (90.3%) | 13 (9.7%) | 1.79 (0.85-3.77, 0.127) |
|  | severe | 6 (66.7%) | 3 (33.3%) | 6.82 (1.95-23.83, 0.003) |
|  | unknown | 33 (89.2%) | 4 (10.8%) | unknown |
| Symptoms course | improving | 253 (95.8%) | 11 (4.2%) | reference |
|  | stable | 143 (92.3%) | 12 (7.7%) | 1.84 (0.81-4.17, 0.145) |
|  | worsening | 24 (77.4%) | 7 (22.6%) | 5.43 (2.10-14.03, <0.001) |
|  | unknown | 41 (89.1%) | 5 (10.9%) | unknown |
| Tier | 1 | 234 (98.7%) | 3 (1.3%) | reference |
|  | 2 | 170 (91.9%) | 15 (8.1%) | 5.29 (1.53-18.32, 0.009) |
|  | 3 | 57 (77.0%) | 17 (23.0%) | 16.24 (4.74-55.59, <0.001) |

### Multivariable analysis

The final model that predicts hospitalization among outpatients in VOMC as shown in table 3 includes (1) risk tier, (2) reported obesity, (3) age ≥ 60, and (4) gender as strata. This model had an overall fit that was statistically significant (p value <0.001). In checking the proportionality of hazards assumption, gender was found to be a time-varying covariate (eAppendix 3). As a result we analyzed gender as strata.^15^ The adjusted hazard ratio for Tiers 2 and 3 compared to Tier 1 were 3.74 (95% CI 1.06-13.27, p=0.041) and 10.87 (95% CI 3.09-38.27, p<0.001), respectively. Age ≥ 60 had an adjusted hazard ratio of 2.53 (95% CI 1.27-5.02, p=0.008) and reported obesity had an adjusted hazard ratio of 2.09 (95% CI 1.06-4.13, p=0.034). Survival curves (Figure 1) show days from symptom onset to hospitalization by Tier. Tier 3 patients were hospitalized earlier and at higher rates for both males and females. At 30 days, 24% of males in Tier 3 were hospitalized versus 9% in Tier 2. For females, 16% in Tier 3 were hospitalized while 6% in Tier 2 were hospitalized at 30 days. Males were hospitalized earlier and more often than females. Males reached the maximum number of admissions for males in our cohort occurred by day 23, while females in our cohort reached maximum hospitalization rates by day 41. Logistic regression performed with the same variables to shadow the Cox regression analysis found similar results with adjusted odds ratios (OR) of 4.87 for Tier 2 and 15.38 for Tier 3 compared to Tier 1. Age ≥ 60 and Reported Obesity both had adjusted ORs similar to their adjusted hazard ratios (table 3). Gender was not statistically significant but was kept in the logistic regression model for comparison to the survival analysis.

**Table 3:** Hazard Ratios and Odds Ratios for Variables with Significant Predictive Value

| Variables | Unadjusted Hazard ratio | Adjusted Hazard ratio*<br>(95% CI, p value) | Adjusted Odds ratio**<br>(95% CI, p value) |
| --- | --- | --- | --- |
| Tier | reference | reference | reference |
| Tier 2 | 5.29 (1.53-18.32, 0.009) | 3.74 (1.06-13.27, 0.041) | 4.87 (1.35-17.57, 0.016) |
| Tier 3 | 16.24 (4.74-55.59, <0.001) | 10.87 (3.09-38.27, <0.001) | 15.38 (4.21-56.20, <0.001) |
| Age ≥ 60 | 3.77 (1.94-7.34, <0.001) | 2.53 (1.27-5.02, 0.008) | 2.94 (1.38-6.25, 0.005) |
| Reported Obesity | 2.27 (1.17-4.41, 0.015) | 2.09 (1.06-4.13, 0.034) | 2.17 (1.01-4.67, 0.048) |
| Male | 1.76 (0.91-3.43, 0.095) | analyzed by strata | 1.94 (0.91-4.18, 0.089) |

\* Cox overall model of fit: Chi square 41.37, $p < 0.001$
\*\* Logistic regression overall model of fit: Chi square 50.79, $p < 0.001$

**Figure 1:**
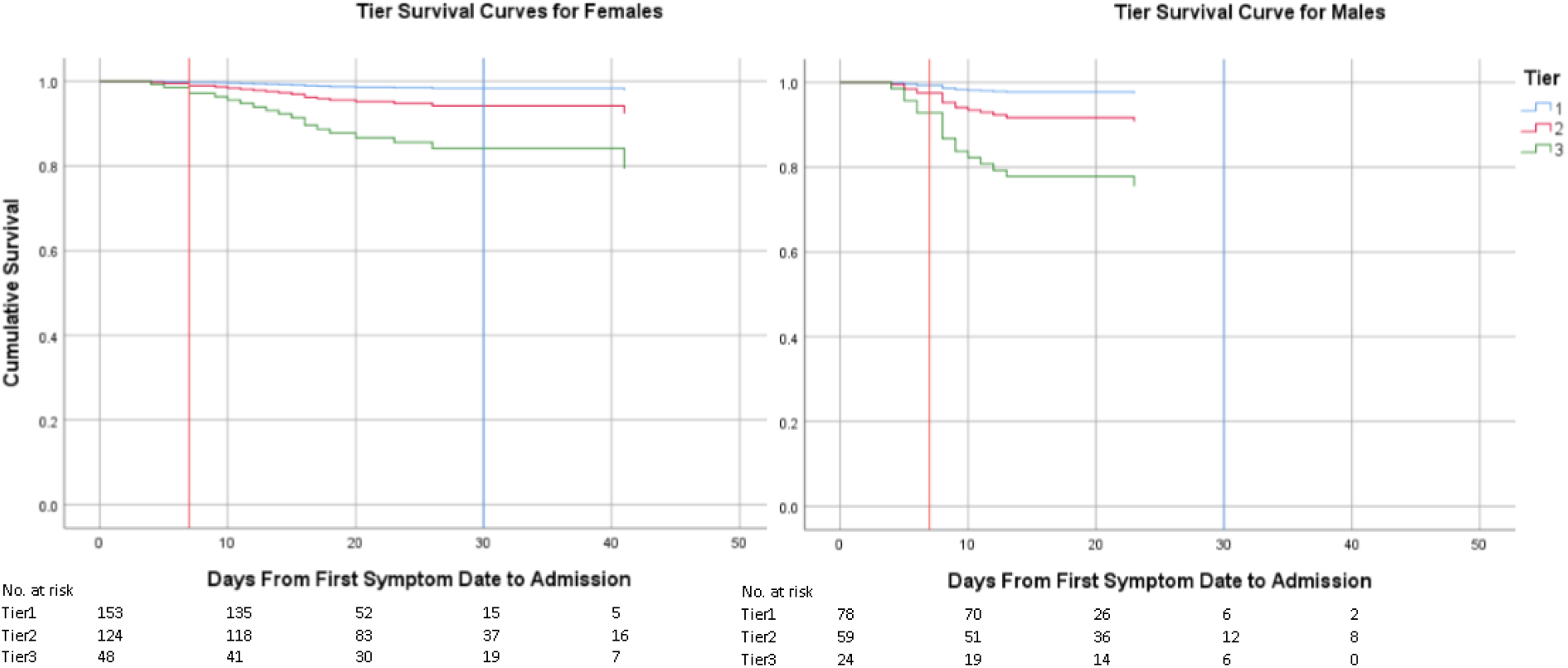
Cox Regression Survival Curves Hospitalization by Risk Tier

### Sensitivity Analysis

We performed sensitivity analysis for obesity to see if using the actual BMI (Corrected Obesity) would be more predictive that reported obesity. The adjusted hazard ratio for corrected obesity was 3.783 (95%CI 1.761-8.126, p< 0.001) with only minor changes in the hazard ratio and p values for Tier and Age ≥ 60 (eAppendix 4).

### Proposed Simplified Tier Model

We looked at factors associated with hospitalization to propose a streamlined risk assessment model to predict patients in the VOMC setting that will not require hospitalization during COVID-19 illness. Table 4 demonstrates the strength of the model using Age<60, no high-risk comorbidities, able to self-isolate, symptom severity mild or none, symptom course stable or improving. This model has no hospitalizations for Proposed Tier 1 patients.

**Table 4:** Proposed Simplified Risk Assessment for Low Risk Patients Tested in Study Cohort

|  |  | Hospital Admission |  |
| --- | --- | --- | --- |
|  |  | No | Yes |
| Tier | 1 | 114 (100.0%) | 0 (0.0%) |
|  | 2&3 | 347 (90.8%) | 35 (9.2%) |

Proposed 4-criteria model for “new Tier 1”: (1) young (age <60), (2) healthy (i.e. no at-risk comorbidities), (3) symptoms mild and stable or improving, (4) able to self-isolate

## Discussion

This study describes the outcomes of outpatients with confirmed COVID-19 who participated in a standardized telemedicine risk assessment and telephone monitoring program. The overall hospitalization rate in this outpatient cohort was 7%, which is lower than other populations reported in the literature.^11^,^16^ While the overall study design is retrospective, our program implemented the risk assessment tool prospectively for all new patients with COVID-19 and we were able to follow all patients until clinical improvement or hospitalization because of the availability of redeployed providers (RNs and APPs), minimizing gaps in data.

### Improving identification of high-risk patients

We found that the risk tier assessment tool predicted hospitalization risk with highly significant results in multivariate analysis and time-to-hospitalization survival analysis. This supports our hypothesis that inclusion of multiple factors in patient assessment (age, risk factors, symptoms, social factors) would most effectively identify absolute hospitalization risk and time to hospital admission. In the same model, age ≥ 60, reported obesity, and male gender are significant predictors of risk which is consistent with previous studies.^10^-^12^ Age and obesity are notable because, despite inclusion in the risk assessment tool, they remain significant even after controlling for tier.

The identification of a small group of outpatients (Tier 3) at highest risk of hospitalization facilitates planning efforts for high-intensity outpatient monitoring with limited follow-up resources and may justify the expanded implementation of the risk assessment tool at the point of care. One question raised by a useful risk-tier rubric is whether it can be codified into a computer-resident algorithm, an artificial intelligence (AI) application. We attempted models with tier as an output rather than input and using the objective and subjective notes and clinical observations as inputs. We were unable to develop such a model, evidently because the tier assignment includes several points where clinical judgment is required and applied. Incorporating that clinical judgment is necessary and is beyond our AI ability at this point.

### Identification of low risk patients

The risk assessment tool identified individuals in Tier 1 at low risk of hospitalization. In our VOMC cohort, this group represented a large volume of the follow-up care. Even with less frequent calls and shorter planned follow-up than other tiers (eAppendix 2), this group received 28.3% of the VOMC follow-up calls (internal data: 1741/6160 calls as of July 2). We were able to simplify criteria for Tier 1 for a proposed “new Tier 1” 4-item risk score that may simplify identification of low-risk individuals. With the end of emergency redeployment and staffing reduction, identification of low-risk individuals allows us to reduce the resources allocated appropriately. As additional remote monitoring tools become available (e.g. automated text message survey) this population may be appropriate to assign “as needed” follow-up instead of proactive monitoring calls.

## Limitations

As a single-center study, the results of our risk assessment validation may not apply to other patient populations. Due to an initial screening strategy which prioritized healthcare workers, we had a high proportion of working-age individuals and relatively few older adults and socially disadvantaged individuals in the study population. This may explain the lower hospitalization rate than other published studies. The time to enrollment in the VOMC (9.3 days) reflects the real-world practice at our clinic, but limits generalizability to acute settings (e.g. urgent care); if same-day results are available, patients may present earlier in illness course.

While the prospective use of risk assessment, minimal gaps in data, and longitudinal assessment are strengths of this study, it is notable that the patients received different levels of observation (frequency of telephone calls, provider type for calls, and duration of follow-up calls) based on assigned tier, which may impact outcomes. A second limitation is the possibility of loss to follow-up: patients could end VOMC care on request and we do not have direct data for outside hospitalizations. However, no missed hospitalizations were identified in a separate quality improvement project involving long-term follow-up calls to 158 discharged VOMC patients.

The risk assessment tool itself has limitations. First, it was designed based on limited data available from early reports of COVID-19 in hospitalized patients and not derived from an outpatient cohort, since none existed at the time. Second, due to the differential risk posed by age ranges and specific comorbidities, the risk tool is relatively complex. The best use was achieved by training a dedicated provider group familiar with its use, but this limits external validity. Even in the optimal setting, we encountered missing data fields (e.g. symptom severity) and underreporting issues (e.g. self-reported obesity vs. BMI). As a related issue, further refinement of Tiers 2 and 3 will require ongoing analysis of risk factors in outpatients.

## Conclusions

Our findings suggest that patients at low risk and high risk for hospitalization may be identified with a telemedicine risk assessment tool incorporating age, medical history, symptom severity, and social factors. The Tier 1 patients in our cohort had low hospitalization rates. We observed increasing odds of hospitalization in Tiers 2 and 3, respectively. External validation of these findings is necessary, but we also recognize that care delivery decisions need to be made immediately in the context of recently escalating cases in the COVID-19 pandemic. It is possible to use these data to create care models targeting highest-risk patients during the highest-risk time periods, but further study of the safety and outcomes of this risk-based approach is needed. This study represents our initial experience with an outpatient telemedicine COVID-19 risk assessment tool. In the absence of clear guidelines on the risk stratification and duration of monitoring of outpatient COVID-19, these data may help guide resource allocation, planning current care structures, and future research.

## Data Availability

Data is not available in de-identified form for distribution at this time.

## Acknowledgements

We would like to acknowledge Dr. Sharon Bergquist, MD and Tina-Ann Thompson, MD for contributions to the risk assessment tool and Dr. David Roberts, MD for the design of the structured intake assessment note. We would also like to acknowledge the members of the Virtual Outpatient Management Clinic including faculty, staff and administrative members of the Paul W. Seavey Comprehensive Internal Medicine Clinic and Emory at Rockbridge Primary Care clinic as well as the physicians, nurses, and advanced practice providers who volunteered from other sites.

## Author Contributions

1. Dr. David Tong and Mr. Taylor had full access to all of the data in the study and take responsibility for the integrity of the data and the accuracy of the data analysis.
2. Concept and design: D. Tong, E. Tong, J. O’Keefe
3. Acquisition, analysis, or interpretation of data: D. Tong, E. Tong, G. O’Keefe, Taylor, J. O’Keefe
4. Drafting of the manuscript: E. Tong, J. O’Keefe
5. Critical revision of the manuscript for important intellectual content: D. Tong, E. Tong, G. O’Keefe, Taylor, J. O’Keefe
6. Statistical analysis: D. Tong, Taylor
7. Obtained funding: None
8. Administrative, technical, or material support: All authors
9. Supervision: D. Tong, E. Tong, J. O’Keefe
10. Conflict of Interest Disclosures: Dr. G. O’Keefe served on an advisory board of Eyepoint Pharmaceuticals in 2019. It is unrelated to the current work.

